# Burden of cardiovascular disease risk factors over 13.5 years in a rural and urban South Indian cohort in comparison with global data

**DOI:** 10.1101/2020.07.03.20145599

**Authors:** Senthil K Vasan, Belavendra Antonisamy, Mahasampath S Gowri, Hepsy Y Selliah, Finney S Geethanjali, Felix Jebasingh, Matthew Johnson, Thomas V Paul, Clive Osmond, Fredrik Karpe, Nihal Thomas, Caroline HD Fall

## Abstract

**Objectives:** To estimate the prevalence, incidence and predictors of cardiovascular disease (CVD) risk factors in the Vellore Birth Cohort, South India.

**Design:** Prospective, cohort study

**Setting:** Population-based cohort of rural and urban communities in and around Vellore city in South India

**Participants:** Non-migrant individuals (n= 962, male 519) were studied at two time points 13.6 years apart i) 1998-2002 (baseline, mean age 28.2 years) and ii) 2013-2014 (follow-up, mean age 41.7 years).

**Main outcome measures:** Prevalence and incidence of CVD risk factors (obesity, central obesity, type 2 diabetes (T2D), hypertension, hypercholesterolemia and hypertriglyceridemia) studied at baseline (1998-2002) and follow-up (2013-2014), prevalence in comparison with the Non-Communicable Disease Risk Collaboration (global) data, incidence in comparison with another Indian cohort from New Delhi (NDBC), and baseline predictors of incident CVD risk factors.

**Results:** The prevalence at 28 and 42 years was 17% and 51% for overweight/obesity, 19% and 59% for central obesity, 3% and 16% for T2D, 2% and 19% for hypertension and 15% and 30% for hypertriglyceridemia. The prevalence of T2D at baseline and follow-up and hypertension at follow-up was comparable with or exceeded that in high income countries despite lower obesity rates. The incidence of most risk factors was lower in Vellore than in the NDBC. Waist circumference strongly predicted incident T2D, hypertension and hypertriglyceridemia.

**Conclusions:** A high prevalence of CVD risk factors was evident at a young age among Indians compared with high and upper-middle income countries, with rural rates catching up with urban estimates. Adiposity predicted higher incident CVD risk, but the prevalence of hypertension and T2D was higher given a relatively low obesity prevalence in global terms. Our findings highlight a high burden of CVD risk factors at younger age with increasing trends observed among rural residents, similar to urban South Indians. Therefore, strategies to prevent CVD should be strengthened in both rural and urban settings to minimise health inequalities and should start young.

**Trial registration:** None

**Key messages:** *What is known on this topic:*

- Cardiovascular disease (CVD) risk burden is increasing in Low- and middle-income countries and contributes significantly to the overall morbidity and mortality.
- Nation-wide data from India demonstrate heterogeneity in the prevalence of CVD risk factors within the country; there is very little incidence data.

*What this study adds:*

- The prevalence of CVD risk factors in India is comparable with or exceeds that in high income countries like USA and Europe, even though obesity levels are lower.
- Adiposity at baseline, particularly waist circumference, is a strong predictor of incident risk factors.
- The prevalence of CVD risk factors is higher in rural than urban communities, but the incidence is comparable or higher in the rural setting indicating that the rural population are catching up

## Introduction

Asian Indians aged 35-64 years lost an estimated 9.2 million years of potentially productive life due to cardiovascular disease (CVD) in 2000, a figure expected to rise to 17.9 million years by 2030.^1^ The Global Burden of Disease (GBD) study reported the age-standardized CVD death rate in India as 282 per 100,000 population in 2017, accounting for 24% of total deaths, which is higher than the global average (233 per 100,000) and high-income countries (HIC) like the United States (151 per 100,000) and United Kingdom (122 per 100,000).^2^

CVD-related mortality in India increased by 34% from 1990 to 2016.^3^ Nationwide representative data on the prevalence of CVD risk factors are available,^4–6^ and the India State-Level Disease Burden Initiative (ISLBDI, part of the GBD study), has recently reported age-standardized prevalence data and trends for diabetes, hypertension and obesity according to the economic transition levels of different Indian states,^7^ and provide useful information on the disease burden. There remains, however, a lack of longitudinal studies examining changes in CVD risk factors over time in the *same* population. Such incidence data, and predictors of incidence, are critical to understanding risk factor dynamics within a population, and to evaluate the effectiveness of preventive interventions.

In the current study, we used longitudinal data from the Vellore Birth Cohort in South India to report trends in the prevalence, incidence, and predictors of key CVD risk factors in a stable, non-migrant rural and urban population followed over 13.6 years from age 28 years in 1998-2002 (baseline) to 42 years in 2013-2014. We compared the prevalence at baseline and follow-up with global rates using data from the Non-Communicable Disease Risk Collaboration (NCDRisC), and compared incidence with another Indian study (the urban New Delhi Birth Cohort study). We also examined 28-year adiposity, rural/urban residence and lifestyle factors as predictors of incident risk factors at 42 years.

## Methods

### Study population

Vellore is situated 135 km from Chennai, the state capital of Tamil Nadu, and has a current population of ∼484,690 individuals. The Vellore Birth Cohort (VBC) includes men and women born in 1969-73 to population-based samples of pregnant women recruited from Vellore town (urban) and adjoining rural villages. Details of the cohort are provided elsewhere.^8^ In brief, singleton newborns (n=10,670) born to mothers in 1969-73, from defined areas of Vellore town and 26 nearby rural villages were followed through birth (Phase-1, n=10,670), infancy (Phase-2, age 0-3 months, n=5,753), childhood (Phase-3, age 6-8 years, n=5,541), and adolescence (Phase-4, age 10-15 years, n=2,672). The adult follow-up included two phases: Phase-5 (1998-2002, mean age 28 years, n=2,181) and Phase-6 (2013-2014, mean age 42 years, n=1,080). The current analysis includes 962 individuals (Rural 457; Urban 505) seen at both adult follow-ups. Participants who migrated between Phases 5 and 6, either rural-urban (n=36) or urban-rural (n=82) were excluded, to avoid any confounding effect of migration.

### Demographic and lifestyle measurements

Data were collected using the same methods at age 28 and 42 years. Educational status, tobacco and alcohol use, physical activity (PA) and socio-economic status (SES) were collected by trained field workers using standardised questionnaires. Attained education was recorded at 7 levels from ‘no schooling’ (level 1, 0 years), primary school (level 2, 1–8 years), middle school (level 3, 9–15 years), high school certificate (level 4, 9–12 years), high school (level 5, >12 years), other graduate (level 6) and professional degree (level 7). Tobacco use was categorised as non-smoker, mild, moderate or heavy smokers and included tobacco chewing and snuff, as well as smoking (indigenous beedi’s and/cigarettes). These were converted into cigarette/beedi equivalents per day and categorized as 0-none, 1-mild (10), 2-moderate (10-20) and 3-heavy (>20). The frequencies and quantity of consumption of beer, wine, and spirits were converted into units of alcohol per week (1 unit=574 ml beer or 125 ml wine or 23 ml spirits). They were categorized as 0=none (0 units), 1=mild (≤7 units), 2=moderate (8–21 units) and 3=heavy (>21 units) or dichotomized into current consumers or non-consumers of alcohol. A possessions score was used to define SES, based on ownership of five household items (electric fan, bicycle, car/jeep, motor bike, television) which was recorded at both time points. PA score was calculated as previously described.^8^

### Anthropometric and biochemical measurements

During both adult phases, anthropometry (height, weight and waist circumference [WC]) was measured using standardised protocols. Blood pressure (BP) was recorded seated using a digital sphygmo-manometer (Omron M3 Corporation, Tokyo, Japan) after five minutes seated. Fasting glucose and lipids and 2-hour blood glucose was measured following a 75g oral glucose tolerance test (OGTT). Glucose was measured on a Roche Cobas 800 Autoanalyser (hexokinase-mediated reaction) excluding participants with known diabetes and lipids by standard enzymatic methods. The study was approved by the institutional ethics committee of the Christian Medical College, Vellore and all participants provided informed consent.

### Endpoints

Continuous variables included anthropometry, systolic and diastolic BP and biochemical markers. Binary endpoints were defined using World Health Organization (WHO) criteria for overweight (BMI ≥25 kg/m^2^), obesity (BMI ≥30 kg/m^2^), central obesity (Men: WC ≥90 cm, Women: ≥80 cm), impaired fasting glucose (IFG, fasting glucose ≥6.1 mmol/l and <7.0 mmol/l), impaired glucose tolerance (IGT, fasting glucose <7.0 mmol/l and 2h-glucose ≥7.8 and <11.1mmol/l), type 2 diabetes (T2D, fasting glucose ≥7.0 mmol/l or 2hOGTT ≥11.1 mmol/l or on anti-diabetic medication) and hypertension (systolic BP [SBP] ≥140 mmHg or diastolic BP [DBP] ≥90 mmHg, or on anti-hypertensives]. National Cholesterol Education Program guidelines were used to define hypertriglyceridemia (serum triglycerides ≥1.5 mmol/l or on lipid-lowering medication), hypercholesterolemia (serum total cholesterol ≥5.2 mmol/l or on medication) and low HDL-cholesterol (HDL-cholesterol Men: <1.03 mmol/l; Women: <1.29 mmol/l or on medication). Incident cases of these binary endpoints were defined as new cases diagnosed at Phase-6 and absent at Phase-5.

### Statistical analysis

Kernel density plots were used to show changes in continuous risk factors between baseline and follow-up. The prevalence of binary endpoints was calculated as the ratio of the total number of cases to the study population at each age and presented as number (percentage). For global comparisons, we acquired age- and year-specific prevalence data (cases per 1000) for obesity,^9^ diabetes,^10^ and hypertension^11^ from NCDRisC collaborators for the 21 most populous countries globally. The VBC prevalence was compared with NCDRisC for the most closely matched age ranges (25-29 years and 40-44 years) and years (2000 and 2013). The incidence in VBC (the proportion of new cases per at-risk sample per year) was compared with a north Indian cohort (New Delhi Birth Cohort [NDBC]) of similar age in which similar CVD risk measurements were made during 1998-2002 and 2006-2009.^12^ Logistic regression was used to identify predictors of incident hypertension, T2D and hypertriglyceridemia, adjusted for age, sex, rural/urban residence, alcohol and tobacco use, socio-economic status, education level, family history and physical activity. We have previously reported the representativeness of cohort members who were followed-up to Phase-5 compared with the remainder of the original birth cohort and showed only small differences in early growth.^13^ In a sensitivity analysis, we assessed the representativeness of the analysis sample by comparing socio-demographic, anthropometric and risk marker data at Phase-5 in those studied and not studied in Phase-6, using t-tests and Chi square tests.

### Patient and Public involvement

Patients were not involved during the study design, conduct and were not consulted to develop patient relevant outcomes or interpret the results. However, results were disseminated through community/cohort meetings and other dissemination plans were discussed. Patients were not invited to contribute to the writing or editing of this document for readability or accuracy.

## Results

The demographic, anthropometric and cardiovascular risk factors of the 962 individuals in Phase-5 and Phase-6 are presented in the **Supplementary Table 1**. Tobacco use decreased among rural men, but remained stable in urban men. Alcohol consumption increased in rural men. Tobacco and alcohol consumption remained non-existent among women in both phases. SES increased and PA decreased in all groups.

**Table 1.** Prevalence of CVD risk factors at Phase-5 (baseline) and Phase-6 (follow-up) in the Vellore Birth Cohort according to sex and place of residence (rural or urban).

| CVD Risk Factor | Phase | Men |  | Women |  |
| --- | --- | --- | --- | --- | --- |
|  |  | Rural<br>(n=240) | Urban<br>(n=279) | Rural<br>(n=217) | Urban<br>(n=226) |
| Overweight/Obese, n (%) | Phase-5 | 16 (6.7) | 53 (18.9) | 20 (9.2) | 72 (31.9) |
|  | Phase-6 | 85 (35.4) | 141 (50.5) | 108 (49.8) | 160 (70.8) |
|  | Change in percentage | +28.7 | +31.6 | +40.6 | +38.9 |
| Obesity, n (%) | Phase-5 | 2 (0.8) | 6 (2.2) | 1 (0.5) | 18 (8.0) |
|  | Phase-6 | 13 (5.4) | 38 (13.6) | 34 (15.7) | 77 (34.1) |
|  | Change in percentage | +4.6 | +11.4 | +15.2 | +26.1 |
| Central obesity, n (%) | Phase-5 | 21 (8.8) | 58 (20.8) | 30 (13.8) | 79 (35.0) |
|  | Phase-6 | 105 (43.8) | 171 (61.3) | 119 (54.8) | 173 (76.6) |
|  | Change in percentage | +35.0 | +40.55 | +41.0 | +41.6 |
| Impaired fasting glucose/<br>Impaired glucose tolerance, n (%) | Phase-5 | 50 (20.8) | 53 (19.0) | 48 (22.1) | 61 (27.0) |
|  | Phase-6 | 28 (11.7) | 41 (14.7) | 33 (15.2) | 51 (23.0) |
|  | Change in percentage | -9.1 | -4.3 | -6.9 | -4.0 |
| Type 2 diabetes, n (%) | Phase-5 | 4 (1.7) | 11 (3.9) | 4 (1.8) | 8 (3.5) |
|  | Phase-6 | 45 (18.8) | 50 (17.9) | 25 (11.5) | 38 (16.8) |
|  | Change in percentage | +17.1 | +14.0 | +9.7 | +13.3 |
| Hypertension, n (%) | Phase 5 | 4 (1.7) | 15 (5.4) | 1 (0.5) | 2 (0.9) |
|  | Phase 6 | 58 (24.2) | 79 (28.4) | 23 (10.6) | 28 (12.4) |
|  | Change in percentage | +22.5 | +23.0 | +10.1 | +11.5 |
| Hypertriglyceridemia, n (%) | Phase-5 | 40 (16.7) | 71 (25.5) | 14 (6.5) | 24 (10.6) |
|  | Phase-6 | 93 (38.8) | 119 (42.7) | 37 (17.1) | 38 (16.8) |
|  | Change in percentage | +22.1 | +17.2 | +10.6 | +6.2 |
| Hypercholesterolemia, n (%) | Phase-5 | 10 (4.2) | 43 (15.4) | 9 (4.2) | 29 (12.8) |
|  | Phase-6 | 38 (15.8) | 76 (27.2) | 19 (8.8) | 42 (18.6) |
|  | Change in percentage | +11.6 | +11.8 | +4.6 | +5.8 |
| Low HDL-cholesterol, n (%) | Phase-5 | 158 (65.8) | 171 (61.3) | 178 (82.0) | 181 (80.1) |
|  | Phase-6 | 110 (45.8) | 153 (54.8) | 162 (74.7) | 157 (69.5) |
|  | Change in percentage | -20.0 | -6.5 | -7.3 | -10.6 |

### Prevalence of CVD risk factors

A consistent increase in the prevalence of binary CVD outcomes was seen (**Table 1)**. Exceptions were IFG/IGT, the prevalence of which decreased, counter-balanced by an increase in T2D, and low HDL-cholesterol. Urban residents had a higher prevalence of these outcomes than rural residents at both time points. However, the increase was comparable in both groups (change in percentage, **Table 1**). The highest change was noticed with central obesity A corresponding increase in mean BMI, WC, BP, and glucose and lipid concentrations was observed from Phase-5 to Phase-6 (**Supplementary Table 1**) with a right shift in distribution (**Figure 1 and Supplementary Figure 1**) for most of the cardiovascular risk factors except fasting glucose.

**Figure 1.**
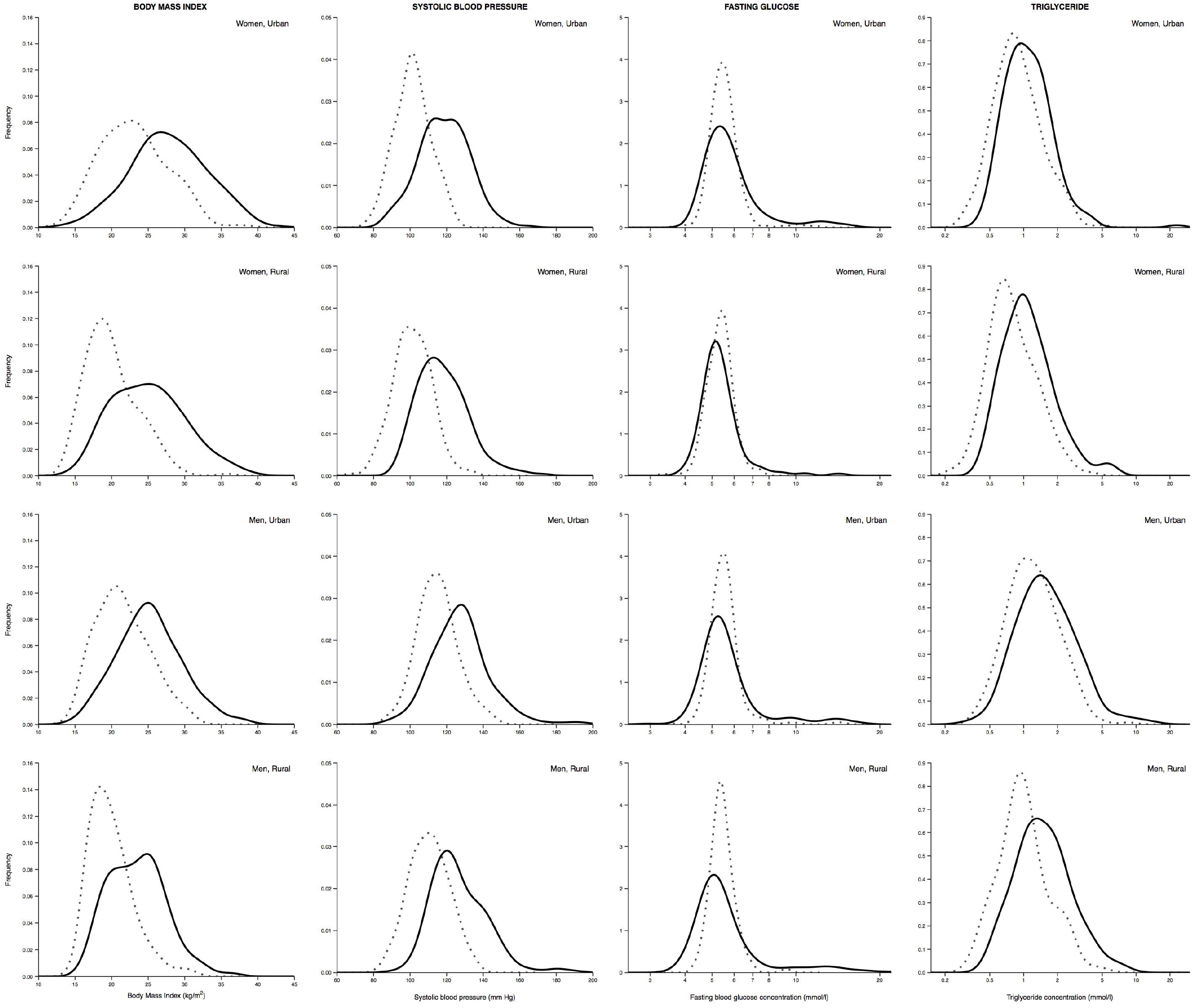
Kernel density plots showing the change in body mass index, systolic blood pressure, fasting glucose and serum triglycerides from Phase-5 to Phase-6 in rural and urban men and women. Dotted lines represent Phase-5 and solid lines represent Phase-6.

### Comparison of prevalence in Vellore with NCDRisC (global) data

**Figure 1 and Supplementary Figure 2** show data from NCDRisC for the prevalence of obesity, T2D and hypertension in 21 countries, with the estimates from Vellore added for comparison. For obesity (**Figure 2**), the countries form two clusters, with a high obesity prevalence at both age/time points in HICs (USA and Europe) and UMICs (upper middle-income countries, Turkey, Mexico, South Africa, Iran and Brazil), and low obesity prevalence in low- (LIC) and low- and middle-income countries (LMICs, Vietnam, Bangladesh and Ethiopia). The NCDRisC estimate for India (a LMIC) falls in the latter group. Exceptions to this pattern are Egypt (a LMIC with high obesity rates) and Japan and South Korea (HICs with low obesity rates). The Vellore estimate is between the two main clusters at both age/timepoints, indicating an obesity prevalence above other LMICs and below UMICs and HICs.

**Figure 2.**
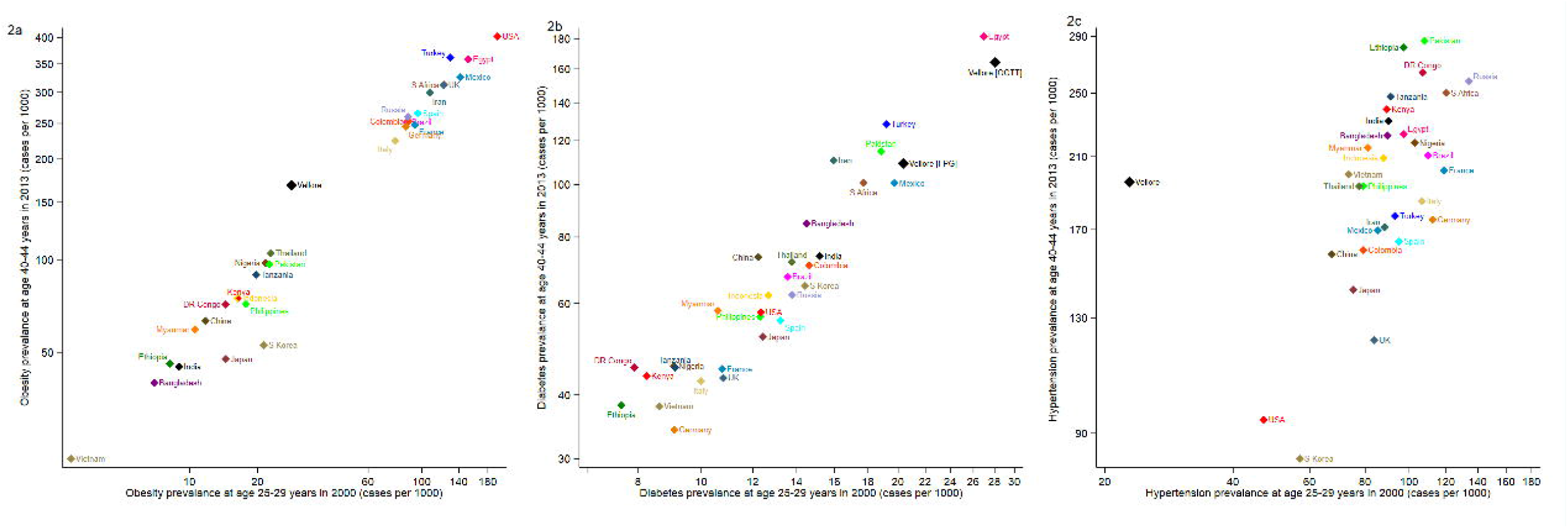
Prevalence (cases per 1000 persons) of obesity, diabetes and diabetes (y axis), in relation to the prevalence of obesity (x axis) at age 40-44 years in 2013 in Vellore compared to global NCDRisC data for the 21 most populous countries at age 25-29 years measured in 2000 versus age 40-44 years measured in 2013 and in Vellore (defined by OGTT) at age 42 years in 2013-2014 (capitalised). OGTT refers to the prevalence estimate based on oral glucose tolerance test criteria (fasting glucose ≥7.0 mmol/l or 2hOGTT ≥11.1 mmol/l) and FPG refers to the prevalence estimate based on fasting plasma glucose only (fasting glucose ≥7.0 mmol/l) in Vellore.

For diabetes (**Figure 2**), values at both age/time points are again correlated, but the ranking of countries differs markedly from that for obesity. HICs (USA, Europe) have a low diabetes prevalence despite high obesity rates. The MICs with comparably high obesity prevalence (Iran, Mexico, Turkey, South Africa, Egypt) have a higher diabetes prevalence than the HICs. Bangladesh, India and Pakistan stand out as countries with low obesity rates but a high diabetes prevalence. Egypt and Vellore stand out with the highest prevalence. The Vellore prevalence is even higher than the NCDRisC estimate for India, and remains strikingly high even when fasting glucose is used to define diabetes, rather than the complete glucose data.

For hypertension (**data not shown**), HICs again had the lowest prevalence and LMICs the highest, with Pakistan and several African countries the leaders. The prevalence doubled in all countries between the two age/time-points. The hypertension prevalence in Vellore at baseline is markedly lower than NCDRisC estimates for all other countries. At follow-up however, the prevalence is comparable to the NCDRisC data for India as a whole and/or other MICs.

### Incidence of CVD risk factors

The highest risk factor incidence was observed for central obesity (**Table 2**). The incidence of low HDL-cholesterol was also high in women. The incidence of obesity markers was higher in the urban than the rural population, but the incidence of other risk factors was similar in both groups. The incidence of all risk factors was 1.5 to 3 times higher in the NDBC compared with the VBC, except that of T2D and low HDL-cholesterol, which was similar in both cohorts.

**Table 2.**
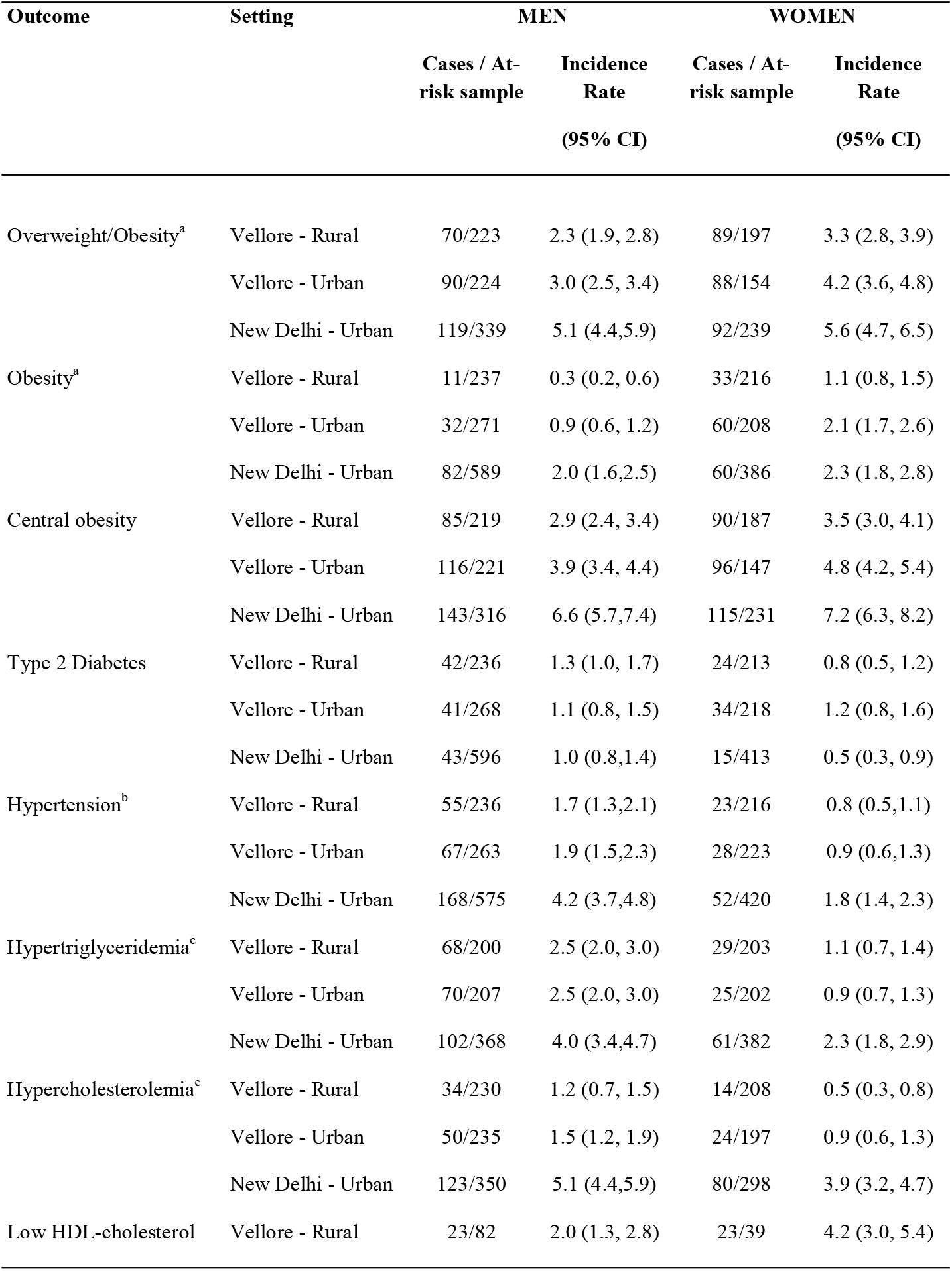
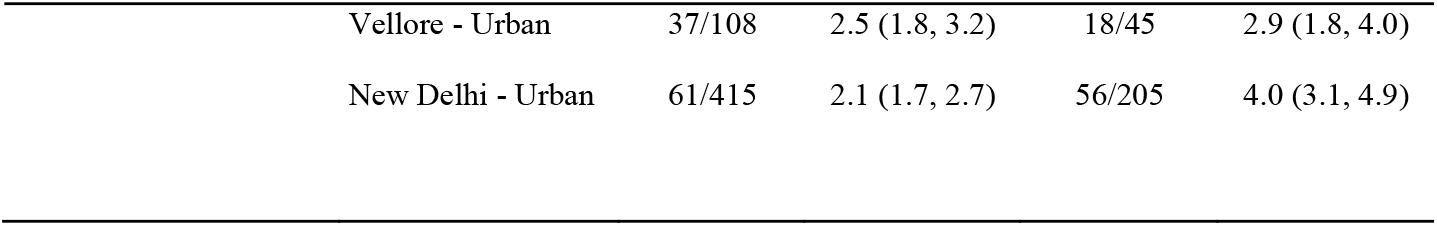
Annual Incidence Rate per 100 persons (95% CI) of CVD risk factors during follow-up according to sex and place of residence in Vellore, and in New Delhi. Missing data: ^a^ BMI status at phase-5 for two study participants (rural men an urban men) and at phase 6 for one study participant (urban men); ^b^ Systolic and diastolic blood pressure values for two participants (1 urban woman and 1 urban man) at phase 6, ^c^ triglyceride and cholesterol levels at phase 6 for one urban man.

### Predictors of incident hypertension, T2D and hypertriglyceridemia

Higher 28-year WC was strong independent predictor of incident T2D, hypertension and hypertriglyceridemia (**Table 3**). Other significant predictors were: (for hypertension) male sex, higher SES, a family history of hypertension, rural residence; (for T2D) rural residence; and (for hypertriglyceridemia) male sex, and alcohol and tobacco use.

**Table 3.** Phase 5 predictors of incident hypertension, type 2 diabetes and hypertriglyceridemia in Phase-6. Model adjusted for Phase-5 predictors, age, sex, place of residence (rural/urban). ^a^p value <0.001; ^b^p value <0.01; ^c^p value <0.05; ^d^Alcohol use and tobacco use categorized as 4 levels from none to heavy; ^e^educational status categorized as 7 levels from illiterate to professional grade;

|  | **Hypertension** | **Type 2 Diabetes** | **Hypertriglyceridemia** |
| --- | --- | --- | --- |
| **Exposures** | **OR (95%CI)** | **OR (95%CI)** | **OR (95%CI)** |
| Gender (Male 0/Female 1) | 0.42 (0.26, 0.66) | 0.88 (0.55, 1.40) | 0.42 (0.29, 0.62) |
| Place of residence (Rural 0/ Urban 1) | 0.63 (0.42, 0.96) | 0.59 (0.39, 0.91) | 0.78 (0.56, 1.09) |
| Age (years) | 1.05 (0.87, 1.27) | 1.06 (0.87, 1.29) | 0.97 (0.83, 1.13) |
| **Phase-5 predictors** |  |  |  |
| Waist circumference (z) | 1.29 (1.07, 1.57) | 2.13 (1.73, 2.62) | 1.52 (1.30, 1.79) |
| Physical activity (logged, z) | 1.15 (0.96, 1.38) | 0.93 (0.77, 1.12) | 1.02 (0.88, 1.18) |
| SES score (z) | 1.36 (1.09, 1.70) | 1.03 (0.82, 1.29) | 1.10 (0.92, 1.31) |
| Alcohol use (0-3)<sup>a</sup> | 1.21 (0.93, 1.59) | 0.96 (0.71, 1.30) | 1.31 (1.05, 1.64) |
| Tobacco use (0-3)<sup>a</sup> | 0.89 (0.64, 1.23) | 1.30 (0.92, 1.84) | 1.35 (1.04, 1.74) |
| Educational status (0-6)<sup>b</sup> | 0.97 (0.85, 1.11) | 1.08 (0.94, 1.24) | 0.97 (0.87, 1.08) |
| Family history (No/Yes) | 1.67 (1.09, 2.56) | 1.13 (0.70, 1.81) | Data not available |

### Sensitivity analysis

For rural women, there were no significant differences in Phase-5 variables between those studied and not-studied in Phase-6 (**Supplementary Tables 2 and 3**). Among rural men there was a small difference in height, and among urban men small differences in age, alcohol intake and physical activity. Urban women studied in Phase-6 had a significantly higher BMI and WC in Phase-5, and slightly higher fasting glucose, total and LDL-cholesterol concentrations than those not studied.

## Discussion

Our results show, in the VBC, that: i) there was a high prevalence of CVD risk factors at age 28 years and a marked increase in prevalence by age 42 years; ii) the prevalence was higher in urban than rural participants at both time points; however, the incidence was similar in both groups and, for diabetes and hypertension it was higher among rural residents; iii) compared with other countries, the prevalence of diabetes and hypertension in Vellore was higher than expected relative to obesity rankings; iv) Higher 28-year WC was independent predictor of incident T2D, hypertension and hypertriglyceridemia.

### High prevalence of CVD risk factors at a young age and low BMI

Asian Indians develop hypertension, T2D, and CVD at a younger age compared with other ethnic groups.^14,15^ Similarly, a high prevalence of these CVD risk factors at relatively low levels of BMI has been recognised for several decades.^16^ Reasons for this remain unclear and could reflect greater genetic/epigenetic susceptibility and/or other causal factors. Genetic studies do not indicate a greater risk of T2D or hypertension in Asian Indians, as risk estimates of single nucleotide polymorphisms are similar to those in Caucasians.^17^ However, evidence to suggest a possible epigenetic mechanism, linked to β-cell dysfunction, is reported.^18^ Traditional lifestyle risk factors operate similarly across different populations and ethnic groups.^19^ The differences in prevalence estimates across different countries apparently exposed to similar lifestyle factors could be explained by early exposure to novel risk factor such as environmental pollutants, ^20^ impaired early growth resulting from foetal and infant early/under nutrition operating at different stages of the life course and the ‘thin-fat’ body composition, inherent of Asian Indians, characterized by a low lean body mass, and a high fat mass distributed more centrally.^21^

### Rural vs urban disease frequency

The rise in CVD risk factors in rural populations (hitherto considered ‘protected’ by lower energy intakes and a higher physical workload) has important public health implications, because 70% of India’s population is rural, with poor access to health care. Recent studies from India and other LMICs have shown a shift in CVD risk burden from urban towards rural areas, probably linked to recent rapid economic development, accompanied by dietary and lifestyle transitions, in rural India.^22,23^ We found a similar increase in the prevalence of all risk factors in the rural and urban populations. The incidence of risk factors in the rural group was comparable to urban rates, and for T2D and hypertension it was higher (**Table 2**). We speculate that the rural population is more sensitive to low adiposity thresholds and other aspects of transition and warrants further investigation.

### Comparison of incidence rates

Incidence data in India are currently limited to three urban studies covering hypertension,^24^ T2D,^25^ and multiple risk factors^12^ and our results, especially the rural data, are an important addition to the literature. The CARRS study of 16,000 men and women aged 20-60 years in Delhi and Chennai found an age-adjusted hypertension incidence over 2 years (2010 to 2012) of 4.7 and 3.6 per 100 per annum respectively.^24^ The CURES study of 2,410 men and women aged ∼50 years in urban Chennai followed over 9 years (2001-2003 to 2012-2013) showed a diabetes incidence of 2.2 per 100 per annum.^25^ The NDBC studied 1,100 men and women, comparable in age to the Vellore cohort, over 6.9 years (1998-2002 to 2006-2009) (**Table 2**). Collectively these studies suggest that the incidence is even higher in large cities than in either rural or urban Vellore possibly related to the differences in the distribution of the specific risk factors in different settings. The differing patterns of disease risk within a country can have important implications in implementing successful measures towards CVD prevention. Non-availability of comparable time-specific incidence data from HICs limits global incidence comparisons.

### Predictors of incident CVD risk factors

Our finding that WC is a strong predictor of incident risk factors is consistent with the known correlations between WC and a range of metabolic abnormalities, including reduced insulin sensitivity and dyslipidemia.^26^ Nearly half our cohort participants were centrally obese in Phase-6, and central obesity showed the greatest increase with age/time of all the adiposity measures (**Table 2**). It has been estimated that 15% and 36% of CVD-and diabetes-related disability-adjusted life-years are attributable to high BMI or central adiposity in Indians.^7^ Biological determinants of CVD risk factors in LMICs are likely to be similar to those of developed countries,^20^ but the drivers of these determinants are likely to differ. Greater SES at baseline predicted incident hypertension, in line with our previous observation of greater levels of CVD risk factors in higher SES groups.^27^ Contrastingly, some recent observations from India have shown higher CVD risk in lower SES strata.^28^ Thus, India may be showing the same systematic shift in CVD risk as that seen in HICs, where CVD appeared first in more affluent social groups and over a period of 50 years became more prominent in deprived social groups.^29^

### Strengths and limitations of the study

Strengths of our study are its prospective design, following the same individuals with the same comprehensive set of measurements at both time points, the use of an oral glucose tolerance test to ascertain diabetes, and contemporaneous rural-urban and global comparisons. The longitudinal nature of the study on same set of non-migrant individuals is first of the kind to provide information on the CVD burden with passage of time. The incidence data is a valuable contribution to currently scarce national and global data from rural populations, and our study was well designed to examine rural-urban differences. A limitation was the narrow age range (42-45 years) of participants, which made it impossible to determine how much of the increased prevalence was due to age and how much to secular trends. There was evidence of possible selection bias among urban women; those who participated in Phase-6 had greater adiposity in Phase-5 than non-participants. Incidence estimates may therefore have been exaggerated in that group.

## Conclusion and policy implications

Our study highlights a high prevalence of CVD risk factors at a young age in India compared with HIC and UMICs, and rapid recent increments which are comparable or even greater, particularly in the rural population. Adiposity is an important driver of this increase, but the prevalence of hypertension and T2D is higher than expected given relatively low obesity rates in global terms, suggesting greater sensitivity to adiposity and/or other unmeasured causal factors, among Indians. CVD risk prevention is one of the important priorities among the nation’s sustainable development goals. Public health strategies aimed at younger age groups and rural populations would curtail the increasing burden CVD burden in India and globally. Results from this study will guide preventive strategies and support policy making.

## Data Availability

All anonymised participant data is available upon request from the corresponding author

## Acknowledgements

We thank the participants and the VBC field team for data collection. We are thankful to Professor Santosh Bhargava and Professor HPS Sachdev for sharing NDBC data. We thank the NCDRisC collaborators for sharing the global age- and year-specific prevalence data.

## Funding/support

Phase 5 follow-up of the study was supported by funds from the British Heart Foundation, UK (BHF_RG/98001 and BHF_CS/15/4/31493). Phase-6 follow-up was supported jointly by the London School of Hygiene & Tropical Medicine, Indian Institute of Public Health, Hyderabad and internal funds from Christian Medical College, Vellore. SKV is funded by British Heart Foundation Clinical Research grant, no. CRM: 0022324.

## Ethical approval

This study was approved by the institutional review board - ethics committee of Christian Medical College, Vellore

## Patient consent

All participants provided written informed consent after a thorough explanation of the research procedures.

## Data sharing

No additional data available

## Supplementary material

**Supplementary Figure 1**

<u>Title:</u> Kernel density plots showing the change in total-, HDL- and LDL-cholesterol from Phase-5 to Phase-6 in rural and urban men and women.

<u>Legend</u> – Dotted lines represent Phase-5 and solid lines represent Phase-6.

**Supplementary Table 1**

<u>Title:</u> Cardiovascular risk factors (continuous variables) at Phase-5 (baseline) and Phase-6 (follow-up) of the Vellore Birth Cohort participants according to sex and place of residence (rural or urban)

<u>Legend:</u> SD – standard deviation; IQR – inter-quartile range

**Supplementary Table 2**

<u>Title:</u> Anthropometry and life style factors in Phase-5: comparison between cohort members studied and not studied in Phase-6

<u>Legend:</u> ^a^p-values from independent-t-test;

^b^p-values from chi-square statistics;

^c^p-values from t-test after log transformation SD – standard deviation; IQR – inter-quartile range

S**upplementary Table 3**

<u>Title:</u> Cardiovascular risk factors in Phase-5: comparison between cohort members studied and not studied in Phase-6

## Notes

### Competing Interest Statement

The authors have declared no competing interest.

### Clinical Trial

Data was used from two cross-sectional time points of a longitudinal cohort

### Author Declarations

The study was approved by the institutional ethics committee of the Christian Medical College, Vellore and all participants provided informed consent.

